# Susceptibility vessel sign as a predictor for recanalization and clinical outcome in acute ischaemic stroke: A Systematic Review and Meta-analysis

**DOI:** 10.1101/2021.01.14.21249827

**Authors:** Si Zhao Tang, Jon Sen

## Abstract

T2^*^-weighted MRI using GRE and SWI sequences can potentially prognosticate the recanalization rate and clinical outcomes in patients with acute ischaemic stroke, using susceptibility vessel sign (SVS) and prominent hypointense vessel sign (PHVS).

A literature search on PubMed, EMBASE databases and other sources from inception up to 01 February 2020 was conducted. 15 studies which reported SVS and PHVS were included in qualitative synthesis. 9 studies on SVS were included in quantitative synthesis i.e. meta-analysis.

Meta-analysis did not show any significant difference in the recanalization rate between SVS (+) group and SVS (-) group (RR = 0.95, 95% CI = 0.87–1.05, p = 0.33). Treatment subgroup analysis (intravenous thrombolysis, IVT- or mechanical thrombectomy, MT-only) does not show significant association between the SVS and IVT-only (RR = 0.73, 95% CI = 0.51-1.05, P=0.09); or MT-only groups (RR = 0.99, 95% CI = 0.90-1.09, P=0.90). No significant association between poor clinical outcome at 3 months and presence of SVS (RR = 1.42, 95% CI = 0.79–2.57, p = 0.24). Treatment subgroup analysis revealed significant association of the SVS and poor clinical outcome at 3 months in the MT-only (RR = 0.67, 95% CI = 0.55–0.82, p = 0.0001) or no thrombolytic treatment (RR = 2.83, 95% CI = 1.69-4.75, p < 0.0001).

In conclusion, there is a significant association between the presence of SVS and poor clinical outcome in patients who underwent MT or without treatment, and no definitive association between the presence of SVS and recanalization rate for acute ischemic stroke.

## Introduction

In recent years, there have been a multitude of studies which investigated the association between gradient-recalled echo (GRE)/susceptibility-weighted imaging (SWI) magnetic resonance imaging (MRI) sequences as a prognostication test for recanalization rate and clinical outcome. 1 2

### Principles of GRE and SWI

The GRE and SWI sequences are T2*-weighted sequences, based on T2* relaxation i.e. the rate of decay in transverse magnetization. The magnetic field inhomogeneities from susceptibility differences between tissues and materials results in accelerated T2* relaxation, leading to the loss of signal intensity on GRE sequence. These magnetic field inhomogeneities cause geometric distortion and artefacts from loss of signal intensity, which are visualised as dark (or hypointense) areas on MR images and sometimes termed as susceptibility artefacts. Magnetic field inhomogeneity can be caused by blood products, paramagnetic contrast agents, iron deposits, air-tissue interfaces and metallic implants. 3

The nomenclature for GRE imaging can be confusing due to variations in sequence design depending on the machine vendors. For the purpose of clarification, the GRE sequence in this article refers to the two-dimensional (2D) GRE sequences, as SWI is also based on GRE imaging but the acquisition is done in a three-dimensional (3D) planes. The GRE sequences are more sensitive compared to SWI sequences to the deleterious T2* effects from the air-tissue interfaces or metallic implants. This is due to the increased thickness of the sections acquired which is generally associated with larger voxel sizes in GRE sequences. 3

SWI is a more recently developed 3D sequence (in relative to GRE) which uses a T2*-weighted technique that exploits the susceptibility differences in magnetic fields together with both magnitude and additional phase information. The phase information is acquired based on local susceptibility differences of the tissues, which then can be used to differentiate paramagnetic substances (such as deoxyhaemoglobin, haemosiderin, ferritin) or diamagnetic substances (such as calcium), depending on the algorithm used in the MRI machine which is vendor-specific. Phase images are more sensitive to magnetic field inhomogeneity in different tissues components as well and provide a superior image contrast in comparison to the GRE sequence. Hence, SWI is considered to be more sensitive in detecting paramagnetic substances i.e. deoxyhaemoglobin. 3

Both GRE and SWI sequences are useful in depicting paramagnetic blood products such as deoxyhaemoglobin, methaemoglobin or hemosiderin within tissues. This is useful in pathological conditions such as cerebral haemorrhage, vascular malformation, haemorrhage in tumour, thrombosed aneurysm et cetera.

### Imaging Markers based on GRE/SWI for Prognostication in Acute Ischaemic Stroke

Several studies have identified and described the usage of this susceptibility sign (i.e. increased in hypointense area due to signal loss) on T2*-weighted imaging within the middle cerebral artery (MCA), cortical and deep cerebral veins as imaging markers 4-8, which are termed as the susceptibility vessel sign (SVS) and prominent hypointense vessel sign (PHVS) in this review article.

The SVS was defined as a hypointense signal that exceeds the diameter of the contralateral artery located at the site of the thrombus by Flacke S. et al. 9 and Rovira A. et al. 10 The postulated mechanism of SVS is due to the presence of deoxygenated haemoglobin in acute clots which is paramagnetic in nature, disrupting the magnetic field homogeneity and causing a rapid dephasing of spins with dramatic signal loss. 11 The SVS sign was more frequently observed in patients with erythrocyte-dominant clots and stroke aetiology of cardiogenic embolus (CE), rather than fibrin-dominant clot and patients with large artery atherosclerosis (LAA). 12 13 This association was further supported by a systematic review by Liu M. et al. 2

The prominent hypointense vessel sign (PHVS) was defined as abnormal hypointensity of the cerebral veins after arterial occlusion, which was initially reported and further subdivided into cortical vessel sign (CVS) and brush sign (BS) by Morita N. et al. 14 The CVS was defined as increased hypointensity and enlargement of vessels along the course of the cerebral cortical veins at the ischemic site compared to the contralateral side, whereas the BS was defined as similar changes seen in unilateral deep white matter subependymal and medullary veins. 14 T2^*^-weighted-based imaging is useful for evaluating venous changes in the affected hemisphere after acute ischaemic stroke due to its extreme sensitivity to paramagnetic substances such as deoxyhaemoglobin, thus reflecting the oxygen extraction fraction and metabolic rate of the affected tissue. 15 In the ischaemic brain, increased oxygen extraction fraction and slow flow contribute to a higher level of deoxyhaemoglobin and venous dilatation, which increases the conspicuity of vessels on SWI. 16 This was proposed to indicate increased oxygen extraction, which correlates well with increased venous and capillary deoxyhaemoglobin levels. 17

There are some more recent studies which either combined both CVS and BS into PHVS topographically 18-20 or using novel quantification method. 21 A study was done by Zhang X et al. in 2017 further anatomically mapped and identified the deep cerebral veins in the BS as thalamostriate vein, septal vein, and internal cerebral vein for patients who had acute ischaemic stroke. The difference in hypointensity of the mapped deep cerebral veins in both hemispheres were then evaluated using quantification method, and the association between susceptibility sign in the deep cerebral veins and clinical outcome were reported. 8

Despite the abundance of published studies which evaluated the prognostic value of these two signs on GRE/SWI, the significance of it in determining recanalization rate and clinical outcomes remains debatable. 1 2 Considering the uncertainty, this systematic review was conducted to assess all current evidences on the prognostic value of SVS and PHVS in patients with acute ischemic stroke, and their recanalization status and clinical outcomes.

### Aim and Hypotheses

- To assess all current evidences on the prognostic values of SVS and PVHS for recanalization status and clinical outcomes in patients with acute ischaemic stroke.
- To validate the null hypothesis that SVS is not associated with recanalization status and clinical outcomes in patients with acute ischaemic stroke.

## Methods

The systematic review was conducted according to the guidelines of Preferred Reporting Item for Systematic Reviews and Meta-analysis (PRISMA) and documented using a PRISMA flow diagram, see *Appendix 1* 23.

### Criteria for considering studies for this review

There is no restriction placed on language or time of publication.

The inclusion criteria for studies were as follows: (i) original studies which enrolled subjects diagnosed with acute ischaemic stroke and performed GRE/SWI before treatment for all subjects (ii) reports the specific finding of SVS and/or PHVS, and its association with recanalization rate, and/or functional clinical outcome such as post-treatment modified Rankin scale (mRS).

Exclusion criteria includes: (i) non-original studies e.g. editorial, case report, systematic review, meta- or pooled-analysis, letters to the editors, conference abstract; studies on animals model; basic science studies (ii) duplicated articles or article with overlapping data (iii) study which is limited in appropriate / relevant data.

### Search methods for identification of studies

Potentially relevant studies were identified by systematically searching PubMed, Embase, and the Cochrane Library from inception up to 25 January 2020 with a combination of terms ‘‘susceptibility vessel sign” and ‘‘stroke” and their entry terms. Conference abstract and reference lists of available records identified in the initial publications were also manually searched to avoid omitting relevant researches. The title and abstract of all studies were screened for potential literature that fulfilled the inclusion criteria. Full text versions were subsequently retrieved to review the eligibility to be included.

### Outcome measures

The outcome was recanalization status and poor clinical outcome at 3 months after onset of stroke.

Successful recanalization is defined as an arterial occlusive lesion (AOL) scale 2 and 3, or modified Mori scale 2 and 3 on any radiological image measured within 24 hours after intravenous thrombolysis (IVT), or TICI 2b to 3 after mechanical thrombectomy (MT) 24.

Clinical outcome was defined as poor for modified Rankin score (mRS) of 3–6, and good for mRs≤2.

## Data collection and analysis

### DATA EXTRACTION

The following data were extracted and tabulated from studies into standardized data extraction forms: study design, year, country/region, sample size, type of MR sequences used (GRE or SWI) and their interpretation, type of reperfusion treatment conducted, recanalization status assessment, functional clinical outcome.

### QUALITY ASSESSMENT

The quality of the included studies was assessed using Quality Assessment of Diagnostic Accuracy Studies-2 i.e. QUADAS-2 25.

### STATISTICAL ANALYSIS

Risk ratio (RR) and 95% confidence interval (CI) were used to determine the associations between positive GRE / SWI findings, recanalization status, functional outcomes, and any influence of treatment. The aim is to compare recanalization and radiological outcomes in patients with characteristic GRE / SVS findings and determine the impact of reperfusion treatment.

The overall effect was tested using z scores calculated by Fisher’s z transformation with significance set at p < 0.05. The chi-squared (χ^2^) and I-squared (I^2^) tests were used to evaluate statistical heterogeneity with significance set at P < 0.10. If there is significant heterogeneity of the studies (I^2^ > 50 or P < 0.10), a random-effect (DerSimonian-Laird) model was used; otherwise, a fixed-effect (Mantel-Haenszel) model was chosen. If there was a possibility that the pooled estimates would be confounded by substantial heterogeneity among the studies, the results were not pooled in order to prevent misinterpretation. To evaluate the stability of the pooled results, sensitivity analyses were conducted by changing the analysis model to random-effects model, sequentially removing individual studies and conducting subgroup analysis. Publication bias was evaluated by Egger’s test and qualitative evaluation of funnel plots for asymmetry. All statistical analyses were performed using Review Manager 5.3 (RevMan) 26.

## Results

### Study Selection

*Appendix 1* presents the PRISMA flow diagram for study selection. A total of 1879 records were identified through EMBASSE and MEDLINE, and 17 additional records were identified through other sources. There were 1315 records left after duplicates were removed. 1277 records were excluded based on title and abstract. 33 records were identified to be assessed via a full-text review for eligibility. A total of 22 studies presented a relationship between SVS/PHVS, recanalization status and / or clinical outcomes in patients with acute ischaemic stroke (AIS). Of the 22 studies, 7 studies were further excluded from qualitative analysis due to either inadequate data 19 27 28, using different definition of SVS 29, non-standardized definition of recanalization 4 30 or mRS was evaluated at 6 months 31 (see *Supplementary Table 1*). No studies were considered to be seriously flawed as per the QUADAS-2.

**Table 1.** Studies with SVS.

| Study | Region | Design | Site | Sample size | T2* sequence | Interpretation | Reperfusion therapy | Recanalization assessment | Clinical outcome assessment |
| --- | --- | --- | --- | --- | --- | --- | --- | --- | --- |
| Aoki J, et al (2015) | Japan | Retrospective | Single centre | 161 | GRE | SVS | IVT | Recanalization at 1 hour | mRS score at 3 months |
| Bourcier R, et al (2015) | France | Retrospective | Single centre | 73 | GRE | SVS | MT | Recanalization at end of MT | mRS score at 3 months |
| Bourcier R, et al (2018); ASTER trial | France | Randomized case-control | Multi-centre | 202 | GRE | SVS | MT | Recanalization at end of MT | mRS score at 3 months |
| Bourcier R, et al. (2019) | France | Retrospective | Multi-centre | 217 | GRE | SVS | MT | Recanalization at end of MT | mRS score at 3 months |
| Liu H, et al (2017) | China | Retrospective | Single centre | 116 | SWI | SVS | N/A | N/A | mRS score at 3 months |
| Payabvash S, et al (2017) | USA | Retrospective | Two centres | 213 | SWI | SVS | IVT / MT / none | N/A | mRS score at 3 months |
| Ritzenthaler T, et al (2016) | France | Retrospective | Multi-centre | 50 | GRE | SVS | IVT | Recanalization at 3 hours | N/A |
| Soize S, et al (2015) | France | Retrospective | Multi-centre | 153 | GRE | SVS | MT | Recanalization at end of MT | mRS score at 3 months |
| Yan S, et al (2016) | China | Retrospective | Single centre | 85 | SWI | SVS | IVT | Recanalization status at follow-up MRA 24 hours after IVT | mRS score at 3 months |

15 of studies which reported SVS and PHVS were included in qualitative synthesis (*Table 1 & 2*), and 9 of studies on SVS were included in quantitative synthesis i.e. meta-analysis.

### Study Characteristics

The 15 studies included 1 randomized case-control study 32, 2 prospective cohort studies 5 18 and 12 retrospective studies 6 8 20-22 33-39. The patient populations were from Asia (China, Japan) in 9 studies 5 6 8 18 20-22 35 39, in Europe (France) in 5 studies 32-34 37 38 or North America (USA) in 1 study 36. 9 out of 15 studies evaluated the significance of SVS in either recanalization or clinical outcome or both 6 22 32-38. Whereas the remaining 6 studies investigated the significance of PHVS in different variations (for e.g. CVS, BS, combining both CV and BS, IPTSV which concept was derived from BS), and its relationship with either recanalization rate or clinical outcome or both 5 8 18 20 21 26 39.

In the 9 studies which evaluated SVS, 6 used GRE sequences and 3 used SWI sequences. In terms of treatment strategy for studies in SVS, 4 recruited patients who underwent MT exclusively 32-34 38, 3 included patients treated with IVT solely 6 22 37, 1 study included patients on different treatment strategies 36 and 1 study only included patients who did not had IVT/MT 35. MT was performed with a stent retriever or an aspiration catheter in all enrolled studies. Of the 9 SVS studies, recanalization status was presented in 7 studies, and clinical outcomes were measured in 7 studies. The details of these SVS studies are presented in *Table 1*.

For the 6 studies which evaluated the different variations of PHVS, the definition of PHVS at the ipsilateral side of stroke and its assessing methods were heterogeneous. The different variations of PHVS include cortical vessel sign (CVS) 20 or asymmetrical cortical vessel sign (ACVS) 18 39 which assessed the cortical cerebral veins, brush sign (BS) 5 20 or asymmetrical medullary vessel sign (AMVS) 39 which focused on the medullary and deep white matter cerebral veins, prominent vessel sign (PHVS) which include both cortical and deep cerebral veins 20 21, ipsilateral prominent thalamostriate vein (IPTSV) 8. 4 studies qualitatively assessed the presence of PHVS 5 18 20 39, and two studies assessed PHVS using quantitative method 8 21. The detailed individual characteristics of these PHVS studies are tabulated in Table 2. Meta-analysis for the studies of PHVS was not performed in view of the heterogeneity in PHVS definition and assessment method.

**Table 2.** Studies with PHVS.

| **Study** | **Region** | **Design** | **Site** | **Sample size** | **T2\* sequence** | **Interpretation** | **Reperfusion therapy** | **Recanalization assessment** | **Clinical outcome assessment** |
| --- | --- | --- | --- | --- | --- | --- | --- | --- | --- |
| **Sun W, et al (2014)** | China | Prospective | Single centre | 572 | SWI | ACVS (i.e. CVS) | N/A | N/A | END at 72 hours<br>mRS score at 3 months |
| **Terasawa Y, et al (2014)** | Japan | Prospective | Single centre | 36 | GRE | BS | IVT | Recanalization on follow-up MRA within 24 hours after IVT | mRS score at 3 months |
| **Wang Y, et al (2018)** | China | Retrospective | Single centre | 40 | SWI | PHVS, CVS, BS | IVT / MT | Recanalization on second CTA or end of MT | mRS score at 3 months |
| **Yu J, et al (2017)** | China | Retrospective | Single centre | 124 | SWI | ACVS (i.e. CVS), AMVS (i.e. BS) | N/A | N/A | mRS score at 3 months |
| **Zhang X, et al (2017)** | China | Retrospective | Single centre | 109 | SWI | IPTSV | IVT | Reperfusion status | mRS score at 3 months |
| **Zhao G, et al (2017)** | China | Retrospective | Single centre | 60 | SWI | Quantitative PHVS | IVT | N/A | NIHSS at 1 and 24 hours; mRS score at 3 months |

### Predictive value of SVS on recanalization rate after reperfusion therapy (IVT or MT only)

7 studies revealed a relationship between presence of SVS and recanalization rate after reperfusion therapy. Meta-analysis did not show any significant difference in the recanalization rate between SVS (+) group and SVS (-) group (RR = 0.95, 95% CI = 0.87–1.05, p = 0.33; see Figure 1) with no significant heterogeneity (P = 0.41, I^2^ = 1%).

**Figure 1:**
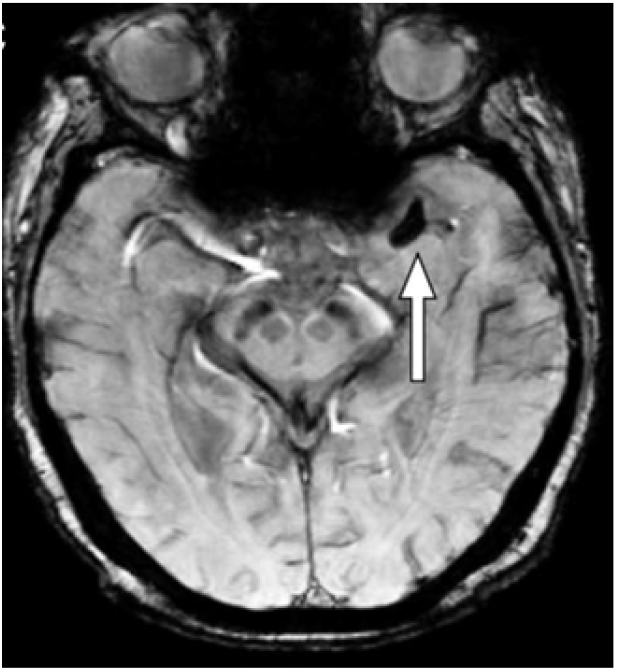
Axial slice MR brain image of a “susceptibility vessel sign” (SVS) on susceptibility-weighted imaging (SWI), which demonstrates hypointense blooming artifact (white arrow) beyond parent vessel lumen in a patient with left middle cerebral artery (MCA) occlusion. Adapted from Yan S et al. 22

In the subgroup analysis with regards to treatment type received (IVT- or MT-only), no significant association is found between the SVS and IVT-only (RR = 0.73, 95% CI = 0.51-1.05, P=0.09) or MT-only groups (RR = 0.99, 95% CI = 0.90-1.09, P=0.90). No significant difference was found between these two treatment subgroups as well (P= 0.11).

### Predictive value of SVS on clinical outcome after 3 months

Pooled analysis was performed for 7 studies which reported the association of SVS and functional outcome using mRS score at 3 months. No significant association between poor clinical outcome at 3 months and presence of SVS (RR = 1.42, 95% CI = 0.79–2.57, p = 0.24; see Figure 2) was demonstrated.

**Figure 2:**
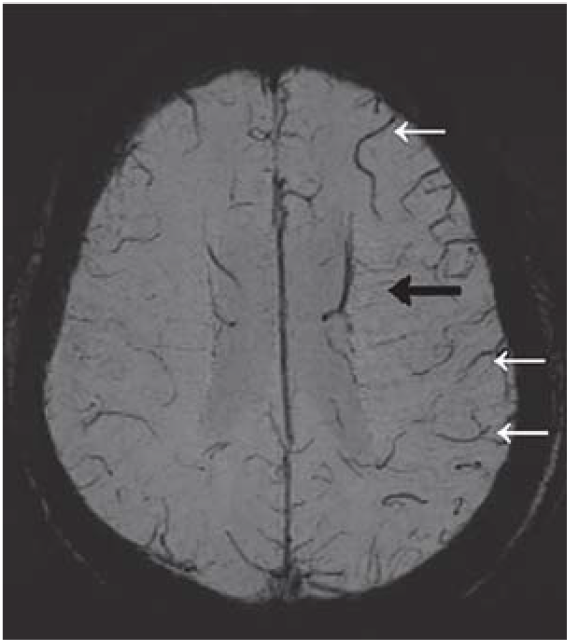
Axial slice MR brain image of a “prominent hypointense vessel sign” (PHVS) on SWI sequence in a patient with left MCA stroke. There are asymmetrical numerous veins with greater hypointense sign and increased diameter in the left frontoparietal cortex and corona radiata. PHVS specifically involving the cortical vessel was defined as “cortical vessel sign” (CVS; white arrows) in some studies, whereas PHVS involving the medullary veins in the deep white matter is described as “brush sign” (BS; black arrow) in some other studies. Adapted from Wang Y et al. 20

**Figure 3:**
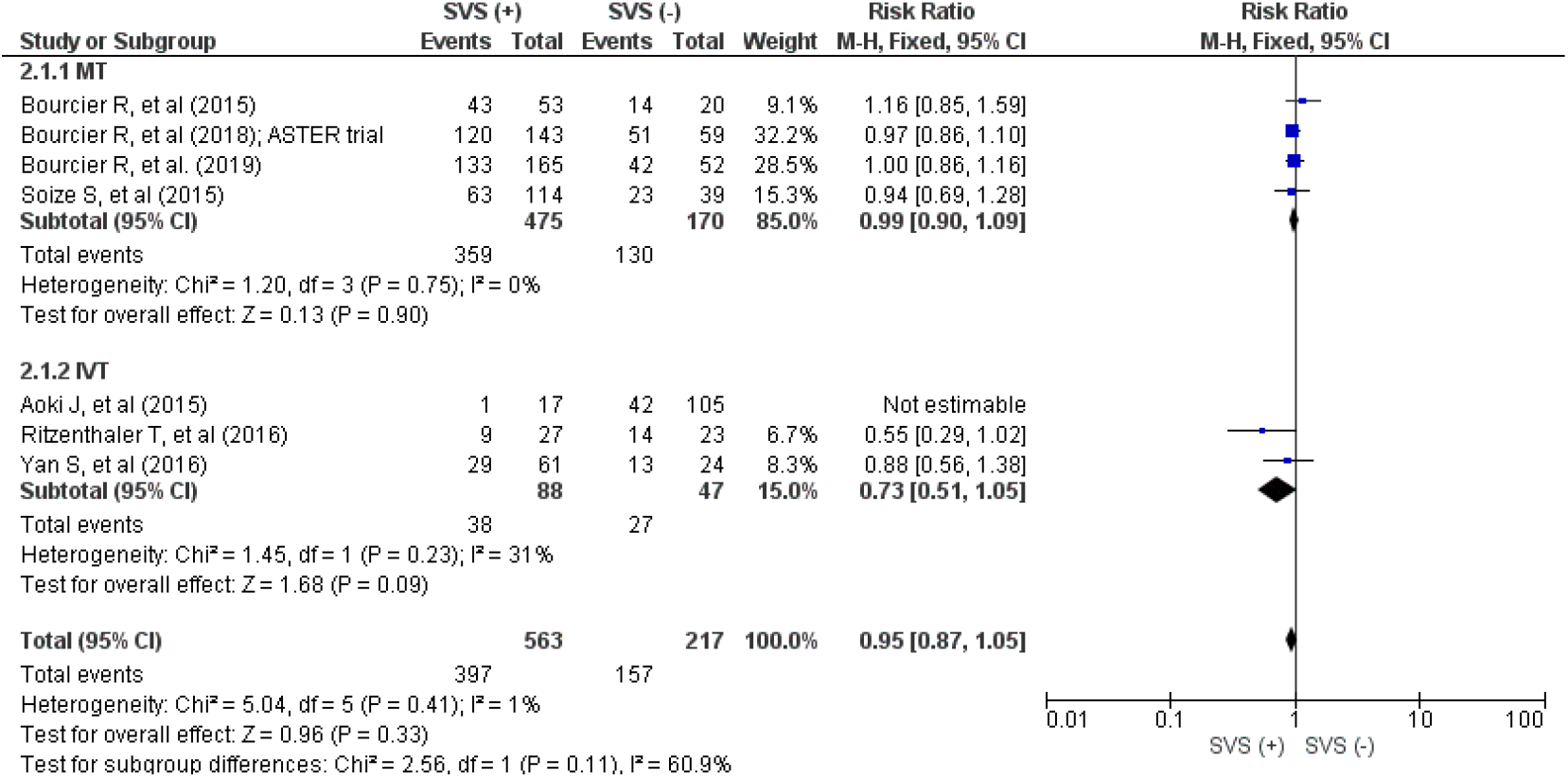
RR and 95% CI values for recanalization rate in SVS versus absence of SVS; fixed-effects model

In treatment subgroup analysis, there is significant heterogeneity in the IVT-only (P = 0.41, I^2^ = 1%) treatment subgroup. Patients with SVS was more likely to achieve poor clinical outcome at 3 months in the MT-only treatment group (RR = 0.67, 95% CI = 0.55–0.82, p = 0.0001), whereas there is no significant association of SVS and poor clinical outcome in the IVT-only group (RR = 1.64, 95% CI = 0.81–3.31, p = 0.17). The group with no thrombolytic therapy given (RR = 2.83, 95% CI = 1.69-4.75, p < 0.0001) and group which received either IVT/MT/no thrombolytic treatment (RR = 7.19, 95% CI = 4.00-12.93, p < 0.0001) both demonstrated significant association between presence of SVS and poor clinical outcome.

**Figure 4:**
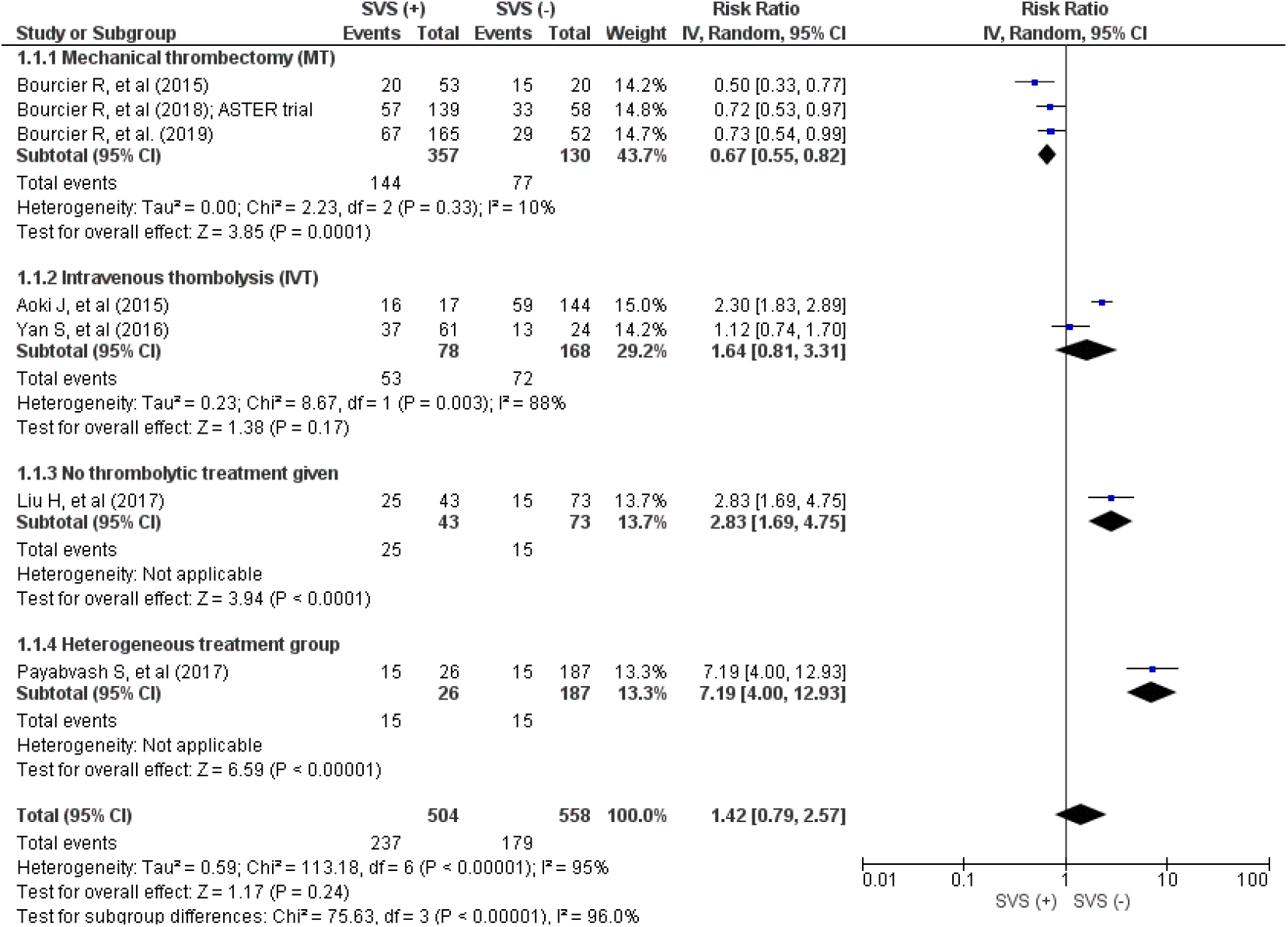
RR and 95% CI values for poor clinical outcome in SVS versus absence of SVS; random-effects model

## Discussion

This review demonstrated prognosticating value of SVS in clinical outcome after acute ischaemic stroke (AIS), and the need of standardizing the different definitions and methods of assessing PHVS and its variations. The pooled estimate based on 7 studies suggested that SVS is not a significant predictor for recanalization outcome after AIS in patients who underwent IVT or MT, and there is no difference between recanalization outcome in patients treated with IVT or MT who had SVS.

### SVS

Despite previous study which reported the presence of SVS as significant predictor for stroke subtype in particularly of cardioembolic origin 2, the presence of SVS as a predictor for recanalization in response to IVT or EVT in AIS patients still remains debatable. It was known that the presence of an SVS in thrombi is related to the composition of a clot. SVS suggests erythrocyte-dominant clot due to its content of predominantly paramagnetic deoxygenated haemoglobin, which interferes with magnetic field homogeneity on susceptibility-weight imaging sequences 13. It was reported that the histopathology of retrieved clots affects the attempts and procedural time of MT for AIS 40, and hence the potential use of SVS to determine its composition and the recanalization outcome in patients who underwent MT. However, this association remains debatable as pooled estimates showed no significant association between SVS and recanalization outcome in patients who underwent MT for AIS.

Pooled estimates also demonstrates significant association between presence of SVS and poor clinical outcome in patients with AIS, but only in two studies which had either patients who had no thrombolytic therapy given due to missed therapeutic window treatment 35, or despite listed as mixed therapeutic strategies but most of the patients (159/213, 74%) had no thrombolytic therapy given 36. Significant association of poor clinical outcome and presence of SVS was also demonstrated with patients who underwent MT-only, but no significant association of poor clinical outcome and presence of SVS is seen in patients who underwent IVT-only. The association of poor clinical outcome and SVS in patients who had no thrombolytic treatment given was expected due to increased recanalization rate, however the association between presence of SVS and poor clinical outcome in patients who underwent MT only is debatable. This highlights the need of more data for further clarification of the predictive value of SVS in clinical outcomes.

### PHVS

The postulated mechanism of blood vessel signal drop on susceptibility-weight imaging in the ipsilateral side of the affected side, i.e. prominent hypointense vessel sign (PHVS) is due to increased oxygen extraction fraction and/or level of deoxyhaemoglobin in the region with hypoperfusion compared to normal brain areas 15-17. In view of the heterogeneity in definition and method of assessment for PHVS, pooled estimate cannot be done to delineate its association with clinical outcome. However, there are two studies in particular which used quantification method to assess this sign, by either comparing the total number of intravenous pixels on the infarction side and on the unaffected side after tracing and automatic segmentation of the shallow and deep veins 21; or specific anatomical subdivided variation of PHVS and reported the association of ipsilateral prominent thalamostriate vein (IPTSV) and poor clinical outcome 8, which their definition and method of assessment appear to be more easily standardized. However only one published study is currently available in each of these two definitions / assessment methods. Further investigation could be directed towards these two signs and their predictive value for clinical outcome after AIS.

### Limitations

There are several limitations to this review. Firstly, most of the included studies were observational studies, which are prone to selection bias. Secondly, there was presence of significant heterogeneity in the meta-analysis for clinical outcome, with source not identified in subgroup analysis. Thirdly, there is lack of standardization of in the description of PHVS. Last but not least, the difference in imaging acquisition technique between GRE and SWI was not accounted, and there is no separation between results based on these sequences.

## Conclusion

In summary, this systematic review and meta-analysis demonstrated that there was significant association between presence of SVS and poor clinical outcome in patients subgroups who underwent MT or no treatment at all; and no definitive association between the presence of SVS and recanalization rate after IVT or MT for acute ischemic stroke. Qualitative systematic review also revealed significant heterogeneity in the definition and method of assessment in PHVS. Given the limitation of current available studies, further large-scale investigations on SVS or predictive value of PHVS / IPTSV on clinical outcomes with common standards and definition of PHVS / IPTSV are required to confirm present conclusions and address some unsolved restriction.

## Data Availability

Literature search on PubMed, EMBASE databases and other sources from inception up to 01 February 2020.

## Appendices

**Appendix 1.**
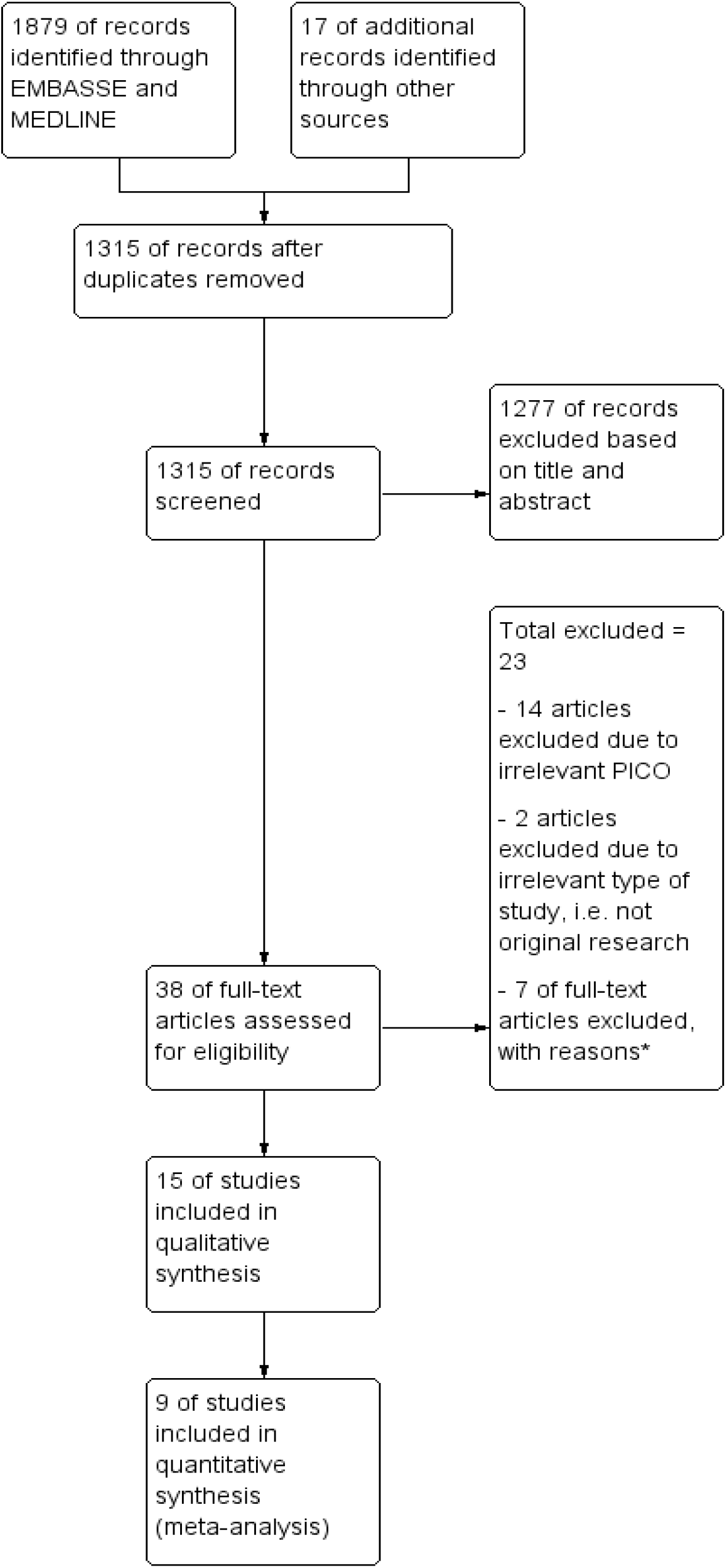
PRISMA flow diagram for SVS/PHVS studies. Adapted from Moher et al., (2009). Preferred Reporting Items for Systematic Reviews and Meta-Analyses: The PRISMA statement.

**Supplementary Table 1:**
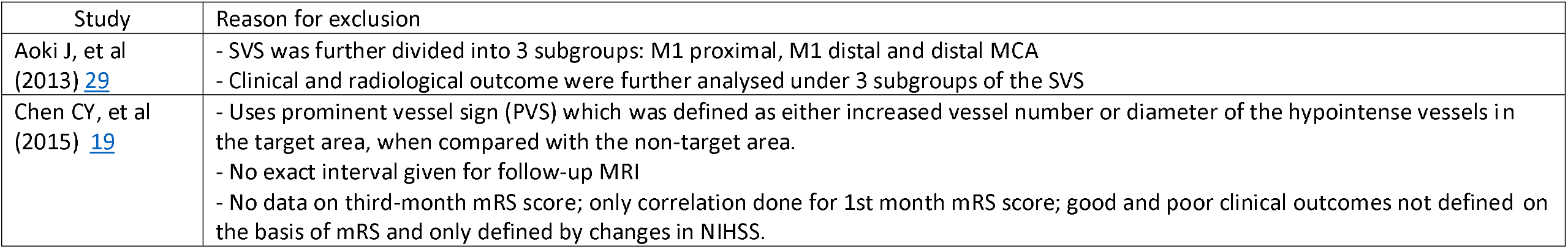
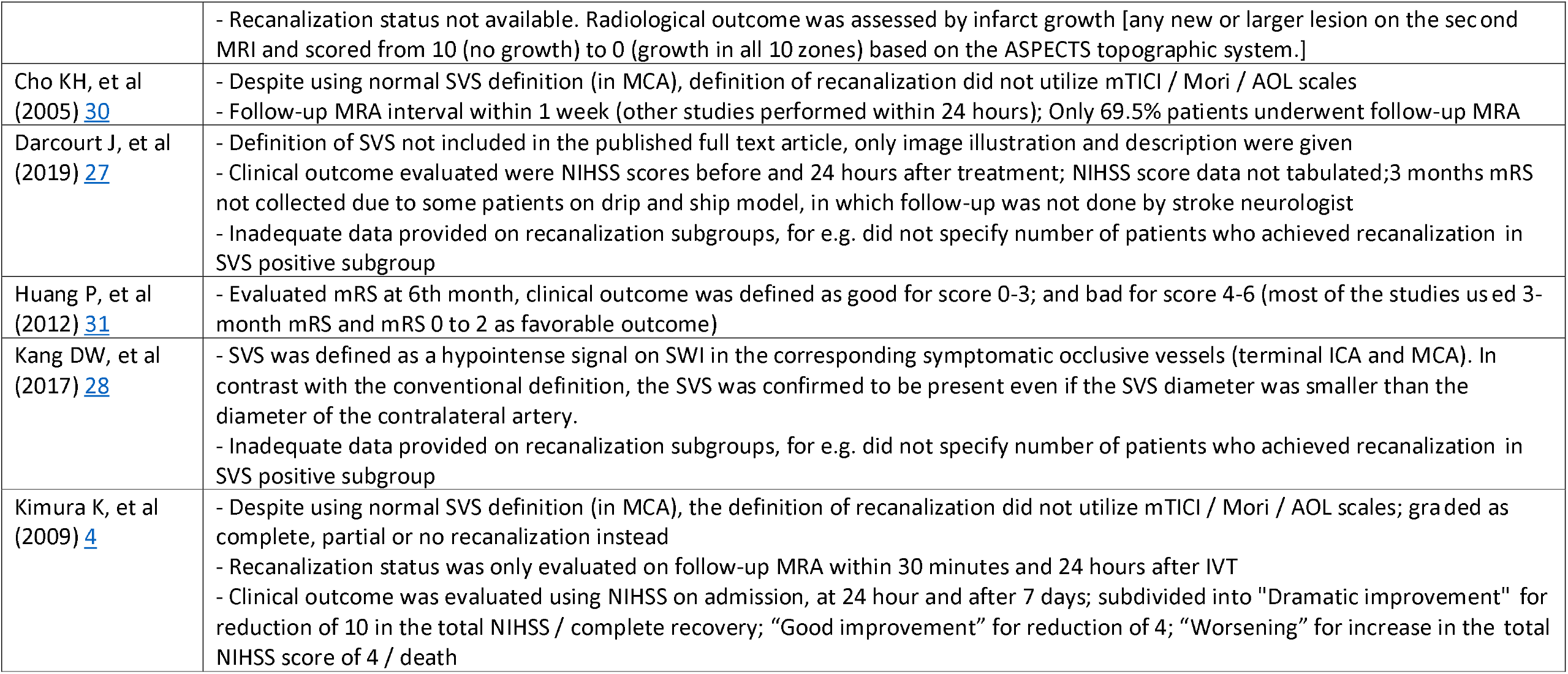
Excluded studies with reasons.

| Study | Reason for exclusion |
| --- | --- |
| Aoki J, et al (2013) 29 | - SVS was further divided into 3 subgroups: M1 proximal, M1 distal and distal MCA<br>- Clinical and radiological outcome were further analysed under 3 subgroups of the SVS |
| Chen CY, et al (2015) 19 | - Uses prominent vessel sign (PVS) which was defined as either increased vessel number or diameter of the hypointense vessels in the target area, when compared with the non-target area.<br>- No exact interval given for follow-up MRI<br>- No data on third-month mRS score; only correlation done for 1st month mRS score; good and poor clinical outcomes not defined on the basis of mRS and only defined by changes in NIHSS. |
|  | - Recanalization status not available. Radiological outcome was assessed by infarct growth [any new or larger lesion on the second MRI and scored from 10 (no growth) to 0 (growth in all 10 zones) based on the ASPECTS topographic system.] |
| Cho KH, et al (2005) 30 | - Despite using normal SVS definition (in MCA), definition of recanalization did not utilize mTICI / Mori / AOL scales<br>- Follow-up MRA interval within 1 week (other studies performed within 24 hours); Only 69.5% patients underwent follow-up MRA |
| Darcourt J, et al (2019) 27 | - Definition of SVS not included in the published full text article, only image illustration and description were given<br>- Clinical outcome evaluated were NIHSS scores before and 24 hours after treatment; NIHSS score data not tabulated; 3 months mRS not collected due to some patients on drip and ship model, in which follow-up was not done by stroke neurologist<br>- Inadequate data provided on recanalization subgroups, for e.g. did not specify number of patients who achieved recanalization in SVS positive subgroup |
| Huang P, et al (2012) 31 | - Evaluated mRS at 6th month, clinical outcome was defined as good for score 0-3; and bad for score 4-6 (most of the studies used 3-month mRS and mRS 0 to 2 as favorable outcome) |
| Kang DW, et al (2017) 28 | - SVS was defined as a hypointense signal on SWI in the corresponding symptomatic occlusive vessels (terminal ICA and MCA). In contrast with the conventional definition, the SVS was confirmed to be present even if the SVS diameter was smaller than the diameter of the contralateral artery.<br>- Inadequate data provided on recanalization subgroups, for e.g. did not specify number of patients who achieved recanalization in SVS positive subgroup |
| Kimura K, et al (2009) 4 | - Despite using normal SVS definition (in MCA), the definition of recanalization did not utilize mTICI / Mori / AOL scales; graded as complete, partial or no recanalization instead<br>- Recanalization status was only evaluated on follow-up MRA within 30 minutes and 24 hours after IVT<br>- Clinical outcome was evaluated using NIHSS on admission, at 24 hour and after 7 days; subdivided into "Dramatic improvement" for reduction of 10 in the total NIHSS / complete recovery; "Good improvement" for reduction of 4; "Worsening" for increase in the total NIHSS score of 4 / death |

